# Factors predicting the visual outcome of intracorneal ring segment for keratoconus

**DOI:** 10.1101/2023.06.22.23291771

**Authors:** Apisit Khanthik, Sasi Yaisawang, Vilavun Puangsricharern, Usanee Reinprayoon, Vannarut Satitpitakul, Ngamjit Kasetsuwan

## Abstract

**Objectives:** To find predictive factors and to construct predictive models using epidemiological and clinical preoperative factors for the visual acuity change after intracorneal ring segment (ICRS) implantation in keratoconus patients.

**Methods:** Medical records of 287 keratoconic eyes implanted with ICRS at Chula Refractive Surgery Centre of a tertiary university hospital (Bangkok, Thailand) between January 2012 and March 2022 were retrospectively reviewed for epidemiological and clinical preoperative variables, including those derived from Scheimpflug tomography. The correlation between these variables and postoperative uncorrected and corrected distance visual acuity change (ΔUDVA and ΔCDVA; logMAR scale) at 6 months was explored. Two hundred forty eight eyes (excluding cases with unmeasurable refraction) were randomized into 2 groups: the equation group (198 eyes) and the validation group (50 eyes). Multiple linear regression analysis was used to develop the predictive model from the equation group.

**Results:** Twenty preoperative variables were statistically correlated with ΔCDVA. Only the preoperative corrected distance visual acuity (CDVAp) strongly correlated with ΔCDVA (R = - 0.746). ΔUDVA correlated with only preoperative visual acuity, corneal astigmatism, and maximum Ambrósio relational thickness (ARTmax). CDVAp, spherical power (SPH), and anterior minimum sagittal curvature (Rmin) were the best predictors of ΔCDVA. The proposed model, ΔCDVA = 0.589 - 0.713×CDVAp - 0.010×SPH - 0.082×Rmin, had an acceptable predictability (R2 = 53.4%). The prediction was correct in 82% of the eyes within 0.22 logMAR.

**Conclusions:** Potential predictive factors and models for ICRS-induced changes in visual acuity were proposed as adjunctive tools for clinicians. Such tools could be used for case selection and during counseling before ICRS implantation to maximize surgical outcomes.

## 1. INTRODUCTION

Keratoconus is an ectatic corneal disorder characterized by progressive biomechanical corneal instability. This can lead to corneal apex thinning, corneal protrusion, irregular astigmatism, and—sometimes—central corneal scarring [1]. Keratoconus is generally bilateral and asymmetrical. Typical onset is during puberty, with progression for around 40 years. Spectacles and rigid contact lenses were the initial treatments used for keratoconus. With progression, this condition can cause irreversible visual loss, leading to corneal transplantation in 10–20% of cases [2]. A reversible and less-invasive surgical option than corneal transplantation is intrastromal corneal ring segment (ICRS) implantation. The ICRS exerts an arc-shortening effect on the corneal architecture, flattening the cornea while improving astigmatism and contact lens tolerance [3–5]. Several studies have emphasized the advantages of ICRS, including its removability, stability, and security, by eliminating the need for an intraocular procedure [6–9].

The corneal response to ICRS in keratoconus remains unpredictable. ICRS improves both uncorrected (UDVA) and corrected visual acuity (CDVA) [9–12]. However, some patients, particularly those whose preoperative corrected visual acuity (CDVAp) was not compromised, experienced worse vision post-implantation [13]. Furthermore, postoperative changes in vision do not regularly correlate with geometric improvements in the cornea [14]. Numerous studies have attempted to determine the relationship between various factors and surgical outcomes to avoid unsatisfactory results [4, 7, 11, 13, 15–26]. However, the impact of these factors on ICRS-induced visual outcomes remains inconclusive, and some factors reportedly increase the risk of post-implantation complications. For example, atopic dermatitis has been associated with increased ICRS extrusion [27].

In this study, we aimed to identify predictors of visual outcomes at 6 months (or within 5 – 12 months) following ICRS implantation. We also created a mathematical model using the abovementioned factors to predict postoperative visual acuities quantitatively. In the future, this model may facilitate patient selection and aid preoperative patient education efforts.

## 2. MATERIALS AND METHODS

### 2.1 Study subjects

This study protocol was approved by the Institutional Review Board (IRB) of the Faculty of Medicine, Chulalongkorn University, Thailand (COA no. 1517/2022 and IRB no. 525/63) and adhered to the tenets of the Declaration of Helsinki. The trial was registered with the Thai Clinical Trial Registry (TCTR20200929001).

This retrospective study was conducted during November 2022 by collecting data from medical records of patients who underwent ICRS implantation at the Chula Refractive Surgery Center, King Chulalongkorn Memorial Hospital, Bangkok, Thailand. Although the authors had access to information that could identify individual patients, the data in the record forms were anonymized to maintain patients’ confidentiality. The primary objective was to identify predictors of postoperative visual acuity change 6 months (or within 5-12 months) following ICRS implantation in patients with keratoconus. The medical records of all eligible cases (351 eyes) were included in this study. The inclusion criteria were as follows: patients with keratoconus implanted with ICRS (Ferrara Ring, AJL, Boecillo, Spain) at the Chula Refractive Surgery Centre between January 2012 and March 2022. Keratoconus diagnosis was based on characteristic signs noted on slit-lamp examination, corneal topography, and tomography. The patient with contact lens intolerance and without contraindication of the ICRS implantation (such as central corneal scar) became a candidate for the surgery. The exclusion criteria were as follows: (i) lack of visual acuity measurement during 5 - 12 months after ICRS implantation (39 eyes), (ii) absence of preoperative data (14 eyes), and (iii) vision-affecting complication and/or ICRS removal within the follow-up period (11 eyes). Ultimately, 64 eyes were excluded from the study, yielding a study cohort of 287 eyes.

### 2.2 Surgical technique

All included subjects underwent ICRS implantation by one of five corneal specialists using femtosecond laser-assisted tunnel creation under topical anesthesia. The arc length, thickness, and number of ring segments were selected according to a previously described nomogram based on the type of keratoconus, the steepest axis, Q-value, and anterior corneal topographic astigmatism. A disposable suction ring was placed and centered after marking the visual axis. Next, an intrastromal tunnel was created by a laser beam with a spot size of 3 µm while focused on a predetermined depth (75% of corneal thickness) from the anterior corneal surface at a 5.0 optical zone. After creating the main incision, the ICRS was implanted using the complete aseptic technique. Postoperative medications included topical steroids and antibiotics.

### 2.3 Data collection and outcome measurement

The epidemiological, pre-implantation, and post-implantation data were obtained from patients’ medical records. The epidemiological data included each patient’s sex, the age at onset, age at ICRS implantation, and history of atopy and eye rubbing. Pre-implantation assessments included uncorrected and corrected distance visual acuities (UDVAp and CDVAp; logMAR), intraocular pressure (IOP; mmHg) by applanation/noncontact tonometer, spherical power (SPH; D) and cylindrical power (CYL; D) from autorefractor, and corneal data evaluated using Scheimpflug tomography (Pentacam HR, Oculus, Wetzlar, Germany). The Scheimpflug tomography-derived variables included the mean keratometry of the front (Km-front; D) and back surfaces (Km-back; D), maximum keratometry (Kmax; D), anterior corneal astigmatism (CA), Q-value of the front (Q-front) and the back surface (Q-back), pachymetry (microns), topographic indices, pachymetric indices, and corneal aberration. Six distances between each pair of four topographic landmarks (apex, pupil center, thinnest point, and Kmax point) calculated by the Pythagorean Theorem were also analyzed. The aberrometry (from Zernike analysis) comprised the total root mean square (total rms), root mean square of lower-order (LOA), and higher-order aberration (HOA). Moreover, the K-factor (KF), which was previously reported as a potential predictor of ΔCDVA, was also determined in this study [21]. The KF was indirectly derived from the Scheimpflug tomography by calculating the multiple products of flattest keratometry and corneal astigmatism of the front corneal surface. The post-implantation visual outcome was assessed by determining changes in the UDVA and CDVA at six months (or within 5-12 months) post-surgery and comparing them to preoperative values (ΔUDVA and ΔCDVA). Holladay’s technique was applied to transform visual acuity into the LogMAR scale for cases where visual acuity was reported as counting fingers, hand motion, light perception, or no light perception. The complications from ICRS implantation were retrieved as secondary outcomes.

### 2.4 Statistical analyses

Statistical analyses were performed using SPSS Statistics for Windows, version 21 (IBM Corporation, Armonk, NY, USA). Considering the large sample size, the data distribution was assumed to be normal. In comparison between the equation group and validation group, Fisher’s exact test and unpaired t-test were used for categorical data and continuous data, respectively. Paired t-test was used to compare pre- and postoperative visual acuity.

Univariate analysis was initially performed to evaluate the correlation between each preoperative variable and the visual outcomes (ΔUDVA and ΔCDVA) using the Pearson correlation coefficient (R). Dummy variables were applied for categorical variables. Female was coded as 0 while male was coded as 1. The history of atopy and eye rubbing was coded as 0 for absence and 1 for presence. The variable with p < 0.2 in this step was considered as a potential predictor and selected for the multivariate regression analysis. The eyes which missed the value of those potential predictors would be excluded from the regression study. In this step, 39 eyes with unmeasurable refractive errors were excluded. The remaining 248 eyes were randomly divided into 2 groups: 198 eyes were assigned to a group for developing the predictive equation (equation group) and the other 50 eyes to a group for validating the proposed equation (validation group).

A stepwise method was applied to construct the predictive model. Model assumptions were evaluated by analyzing the Durbin-Watson test (to confirm the lack of correlation between errors), mean Cook’s distance (to detect influential points or outliers), collinearity tolerance, and the variance inflation factor (VIF). The level of significance was set at *p* < 0.05.

## 3. RESULTS

Among 287 eyes of 230 patients was comprised of 154 (53.7%) right and 133 (46.3%) left eyes. There were 161 males (70.0%) and 69 females (30.0%). The mean age at onset was 20.11 ± 7.86 years (range: 7–50 years), while the mean age at ring implantation time was 26.97 ± 8.56 years (range: 13–59 years). There were 149 (51.9%) eyes with atopy and 177 (61.7%) with frequent eye rubbing. Table 1 summarizes the preoperative numerical data’s means, standard deviations, and ranges. The mean duration of postoperative measurement was 7.0 ± 1.4 months (5.0–12.0 months). We observed significant improvement in the UDVA and CDVA from 1.08 ± 0.55 logMAR to 0.73 ± 0.44 logMAR (Snellen equivalent from 20/239 to 20/109) and 0.51 ± 0.42 logMAR to 0.35 ± 0.28 logMAR (Snellen equivalent from 20/65 to 20/44), respectively. The means of ΔUDVA and ΔCDVA were -0.34 ± 0.48 (95% CI: - 0.40 to -0.29, *p* < 0.001) and -0.17 ± 0.35 (95% CI: -0.21 to -0.13, *p* < 0.001), respectively.

**Table 1.** Mean, standard deviation, minimum, and maximum of the preoperative variables (n = 287 eyes)

| Variables | Mean ± SD | Min | Max | Variables | Mean ± SD | Min | Max |
| --- | --- | --- | --- | --- | --- | --- | --- |
| UDVAp, log MAR | 1.08 ± 0.55 | -0.04 | 2.30 | DAT, mm | 0.72 ± 0.42 | 0.10 | 2.94 |
| CDVAp, log MAR | 0.51 ± 0.42 | -0.04 | 2.30 | DAP, mm | 0.44 ± 0.25 | 0.01 | 1.35 |
| IOP*, mmHg | 10.1 ± 2.7 | 3.0 | 19.0 | DAK, mm | 0.90 ± 0.85 | 0.06 | 4.41 |
| SPH**, D | -6.30 ± 5.10 | -27.00 | 2.37 | DTK, mm | 0.95 ± 0.64 | 0.14 | 4.84 |
| CYL**, D | -5.99 ± 2.97 | -14.00 | 0.00 | DTP, mm | 1.04 ± 0.49 | 0.13 | 3.28 |
| Km-front, D | 51.65 ± 5.43 | 40.10 | 68.80 | DPK, mm | 1.25 ± 0.98 | 0.09 | 4.71 |
| Km-back, D | -7.80 ± 1.06 | -11.35 | -5.35 | ISV | 103.2 ± 41.1 | 21.0 | 346.0 |
| K Factor | 344.9 ± 193.5 | 6.1 | 1055.6 | IVA | 0.88 ± 0.51 | 0.07 | 4.15 |
| Kmax, D | 62.4 ± 8.8 | 47.2 | 88.1 | KI | 1.25 ± 0.16 | 0.88 | 2.30 |
| CA, D | 6.13 ± 3.07 | 0.10 | 18.20 | CKI | 1.10 ± 0.07 | 0.97 | 1.34 |
| Q-front | -1.20 ± 0.60 | -6.30 | 0.01 | IHA | 41.0 ± 31.5 | 0.1 | 158.3 |
| Q-back | -1.12 ± 0.54 | -2.45 | 0.30 | IHD | 0.13 ± 0.08 | 0.00 | 0.48 |
| CPT, μm | 471.1 ± 40.6 | 328.0 | 595.0 | Rmin | 5.51 ± 0.74 | 3.83 | 7.15 |
| CAT, μm | 457.6 ± 44.6 | 295.0 | 596.0 | I-S value | 5.38 ± 4.86 | -12.98 | 32.33 |
| CTT, μm | 447.2 ± 44.4 | 285.0 | 588.0 | total rms | 12.85 ± 6.15 | 2.42 | 48.13 |
| BADD | 11.18 ± 5.11 | 1.46 | 27.09 | LOA, rms | 12.45 ± 5.96 | 2.39 | 46.32 |
| Plavg | 2.48 ± 1.07 | 0.62 | 8.08 | HOA, rms | 3.09 ± 1.63 | 0.33 | 13.08 |
| ARTmax | 149.5 ± 74.1 | 27.0 | 517.0 | | | | |

\*number of subjects = 229
\*\*number of subjects = 248
**Abbreviations:** IOP, intraocular pressure; UDVAp, preoperative uncorrected distance visual acuity; CDVAp, preoperative corrected distance visual acuity; SPH, spherical power; CYL, astigmatism; Km-front, mean keratometry of front surface; Km-back, mean keratometry of back surface; KF, K factor; Kmax, maximum keratometry; CA, anterior corneal astigmatism; Q-front, Q-value of front surface; Q-back, Q-value of back surface; CPT, thickness at the pupil center; CAT, thickness at the apex; CTT, thickness at the thinnest point; BAD D, Belin/Ambrósio enhanced ectasia display; Plavg, average pachymetric progression index; ARTmax, Ambrósio relational thickness maximum; DAT: distance from apex to thinnest point; DAP: distance from apex to pupil center; DAK: distance from apex to maximum keratometry point; DTK: distance from thinnest point to maximum keratometry point; DTP: distance from thinnest point to pupil center; DPK: distance from pupil center to maximum keratometry point; ISV, index of surface variance; IVA, index of vertical asymmetry; KI, keratoconus index; CKI, central keratoconus index; IHA, index of height asymmetry; IHD, index of height decentration; Rmin, anterior minimum sagittal curvature; rms, root mean square; LOA, corneal lower order aberration; HOA, corneal higher order aberration.

The patient demographic data of the equation group and validation group were presented in Table 2. showing the similarity between these two groups.

**Table 2.** Patient demographics and clinical data of equation group and validation group

| Variables | Equation group<br>(n= 198) | Validation group<br>(n = 50) | p |
| --- | --- | --- | --- |
| Sex (male, female) | 125, 73 | 37, 13 | 0.10* |
| Age at onset, yr | 20.73 ± 7.93 | 19.44 ± 8.62 | 0.315† |
| Age at OR, yr | 27.50 ± 8.08 | 26.96 ± 10.30 | 0.693† |
| UDVAp, logMAR | 1.10 ± 0.56 | 0.98 ± 0.51 | 0.170† |
| CDVAp, logMAR | 0.47 ± 0.40 | 0.53 ± 0.38 | 0.315† |
| Spherical power, D | -6.41 ± 5.30 | -5.84 ± 4.22 | 0.481† |
| Cylindrical power, DD | -5.89 ± 3.02 | -6.38 ± 2.78 | 0.295† |
| Km-front, D | 50.67 ± 4.85 | 51.94 ± 5.54 | 0.109† |
| Km-back, D | -7.62 ± 0.98 | -7.85 ± 1.07 | 0.146† |
| Kmax. D | 60.5 ± 8.0 | 62.77 ± 8.39 | 0.082† |
| Q-front | -1.11 ± 0.61 | -1.24 ± 0.55 | 0.147† |
| Q-back | -1.02 ± 0.52 | -1.19 ± 0.55 | 0.051† |
| CTT, μm | 453.0 ± 43.1 | 442.7 ± 46.6 | 0.069† |
| Total rms | 11.88 ± 5.80 | 13.07 ± 5.69 | 0.097† |
| ΔUDVA, logMAR | -0.37 ± 0.48 | -0.23 ± 0.45 | 0.056† |
| ΔCDVA, logMAR | -0.14 ± 0.34 | -0.15 ± 0.25 | 0.906† |
| Follow-up time | 6.83 ± 1.27 | 7.19 ± 1.39 | 0.087† |

\*Comparisons between two groups were made using Fisher's exact test.
†Comparisons between the two groups were made using unpaired t-tests.
Values are presented as mean ± SD

### 3.1 Univariate linear regression analysis

Table 3 summarizes the Pearson correlation coefficients between the postoperative outcomes (ΔUDVA and ΔCDVA) and each preoperative variable. Neither ΔUDVA nor ΔCDVA significantly correlated with the epidemiological variables. Four variables were significantly correlated with ΔUDVA: UDVAp (R = -0.643, *p* < 0.001), CDVAp (R = -0.337, *p* < 0.001), CA (R = 0.119, p = 0.043) and ARTmax (R = 0.118, *p* = 0.046).

**Table 3.** Pearson correlation between the preoperative variables and the postoperative change in uncorrected/corrected distance visual acuities (n = 287)

| Variables | UDVAchange |  | CDVAchange |  | Variables | UDVAchange |  | CDVAchange |  |
| --- | --- | --- | --- | --- | --- | --- | --- | --- | --- |
|  | R | p | R | p |  | R | p | R | p |
| Sex<sup>a</sup> | 0.111 | 0.06 | 0.028 | 0.635 | BADD | -0.072 | 0.224 | -0.301<sup>b</sup> | <0.001 |
| Age_onset | -0.026 | 0.663 | -0.046 | 0.438 | Plavg | -0.078 | 0.186 | -0.311<sup>b</sup> | <0.001 |
| Age_OR | -0.065 | 0.273 | 0.002 | 0.968 | ARTmax | 0.118<sup>b</sup> | 0.046 | 0.251<sup>b</sup> | <0.001 |
| Atopy<sup>a</sup> | 0.046 | 0.434 | 0.014 | 0.815 | DAT | -0.052 | 0.383 | 0.056 | 0.341 |
| Eye rubbing<sup>a</sup> | 0.006 | 0.916 | 0.034 | 0.563 | DAP | -0.054 | 0.362 | -0.100 | 0.091 |
| UDVAp | -0.643<sup>b</sup> | <0.001 | -0.367<sup>b</sup> | <0.001 | DAK | -0.06 | 0.309 | 0.097 | 0.100 |
| CDVAp | -0.337<sup>b</sup> | <0.001 | -0.746<sup>b</sup> | <0.001 | DTK | -0.109 | 0.064 | 0.053 | 0.371 |
| IOP* | 0.05 | 0.447 | -0.005 | 0.937 | DTP | -0.084 | 0.157 | 0.006 | 0.916 |
| SPH** | 0.056 | 0.381 | 0.123 | 0.052 | DPK | -0.036 | 0.549 | 0.110 | 0.062 |
| CYL** | 0.007 | 0.918 | -0.112 | 0.079 | ISV | -0.045 | 0.452 | -0.215<sup>b</sup> | <0.001 |
| Km-front | -0.085 | 0.152 | -0.344<sup>b</sup> | <0.001 | IVA | -0.058 | 0.331 | -0.034 | 0.567 |
| Km-back | 0.049 | 0.413 | 0.310<sup>b</sup> | <0.001 | KI | -0.057 | 0.336 | -0.109 | 0.066 |
| KF | 0.086 | 0.147 | -0.151<sup>b</sup> | 0.010 | CKI | 0.021 | 0.721 | -0.179<sup>b</sup> | 0.002 |
| Kmax | -0.016 | 0.788 | -0.262<sup>b</sup> | <0.001 | IHA | 0.073 | 0.218 | 0.033 | 0.573 |
| CA | 0.119<sup>b</sup> | 0.043 | -0.079 | 0.182 | IHD | -0.051 | 0.391 | -0.102 | 0.083 |
| Q-front | 0.038 | 0.526 | 0.239<sup>b</sup> | <0.001 | Rmin | -0.004 | 0.941 | 0.239<sup>b</sup> | <0.001 |
| Q-back | -0.004 | 0.947 | 0.236<sup>b</sup> | <0.001 | IS | -0.032 | 0.584 | -0.001 | 0.992 |
| CPT | 0.076 | 0.201 | 0.243<sup>b</sup> | <0.001 | total rms | 0.008 | 0.898 | -0.121<sup>b</sup> | 0.040 |
| CAT | 0.089 | 0.132 | 0.256<sup>b</sup> | <0.001 | LOA | 0.012 | 0.843 | -0.117<sup>b</sup> | 0.048 |
| CTT | 0.104 | 0.079 | 0.284<sup>b</sup> | <0.001 | HOA | -0.055 | 0.352 | -0.163<sup>b</sup> | 0.006 |

<sup>a</sup> All dummy variables are coded as 0 for “absent” or 1 for “present” (except for sex: 0 = female, 1 = male).
<sup>b</sup> Correlation is significant at the 0.05 level (2-tailed).
\*Number of subjects = 229
\*\*number of subjects = 248
**Abbreviations:** R, Pearson correlation coefficient; ΔUDVA and ΔCDVA, change in uncorrected/corrected distance visual acuities at 6 months after the surgery; OR, operation date (intracorneal ring insertion); IOP, intraocular pressure; UDVAp, preoperative uncorrected distance visual acuity; CDVAp, preoperative corrected distance visual acuity; SPH, spherical power; AST, astigmatism; Km-front, mean keratometry of front surface; Km-back, mean keratometry of back surface; KF, K factor; Kmax, maximum keratometry; CA, anterior corneal astigmatism; Q-front, Q-value of front surface; Q-back, Q-value of back surface; CPT, thickness at the pupil center; CAT, thickness at the apex; CTT, thickness at the thinnest point; BAD D, Belin/Ambrósio enhanced ectasia display; Plavg, average pachymetric progression index; ARTmax, Ambrósio relational thickness maximum; ISV, index of surface variance; IVA, index of vertical asymmetry; KI, keratoconus index; CKI, central keratoconus index; IHA, index of height asymmetry; IHD, index of height decentration; Rmin, anterior minimum sagittal curvature; HOA, higher order aberration.

Twenty preoperative variables were statistically correlated with ΔCDVA. Only CDVAp demonstrated a strong correlation with ΔCDVA (R = -0.746). In contrast, the others had a weak correlation (|R| < 0.40), including UDVAp, Km-front, Km-back, KF, Kmax, Q-front, Q-back, thickness at pupil center (CPT), thickness at the apex (CAT), thinnest pachymetry (CTT), Belin/Ambrósio Enhanced Ectasia Display (BADD), average pachymetric progression index (PIavg), ARTmax, the index of surface variance (ISV), central keratoconus index (CKI), Rmin, total rms, LOA, and HOA.

### 3.2 Multivariable linear regression analysis

#### 3.2.1 Predictive model for ΔUDVA after six months

According to univariate analysis, twelve preoperative variables with p < 0.20 were introduced to the multiple regression analysis. By applying the stepwise method, UDVAp, and PIavg were selected as explanatory variables of ΔUDVA. (summary of the result is shown in Table 4). The proposed equation was

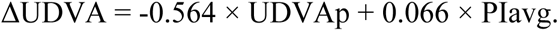

**Table 4.** Stepwise multivariate regression analysis of selected factors to predict the postoperative change of uncorrected distance visual acuity (ΔUDVA) after ICRS implantation

| Variables | Unstandardized Coefficients |  | Standardized Coefficients | p-value | Collinearity Statistics |  |
| --- | --- | --- | --- | --- | --- | --- |
|  | B | 95% CI | Beta |  | Tolerance | VIF |
| (Constant) | 0.094 | -0.061, 0.248 |  | 0.234 |  |  |
| UDVAp | -0.564 | -0.66, -0.468 | -0.659 | <.001 | 0.935 | 1.07 |
| Plavg | 0.066 | 0.01, 0.122 | 0.133 | 0.021 | 0.935 | 1.07 |

$R^2 = 0.408$ ; adjusted $R^2 = 0.402$ ; $F = 67.101$ ( $p < 0.001$ ); mean Cook's distance = $0.0060 \pm 0.014$ ; Durbin-Watson test = 2.079
**Abbreviation:** CI, confidence interval; UDVAp, preoperative uncorrected distance visual acuity; Plavg, average pachymetric progression index; VIF, variance inflation factor

The predictability (R^2^) was 0.408, and the adjusted R^2^ was 0.402, with F = 67.101 (*p* < 0.001). No influential points or outliers were detected (mean Cook’s distance, 0.006 ± 0.014). The independence of the residuals (Durbin-Watson test = 2.079) and the lack of multicollinearity were confirmed.

#### 3.2.2 Predictive model for ΔCDVA after six months

Twenty-eight preoperative variables with p < 0.20 were recruited for the multiple regression analysis. Using the stepwise method, CDVAp, SPH, and Rmin were selected as explanatory variables of ΔCDVA. (summary of the result is shown in Table 5). The proposed equation was

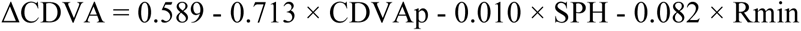

**Table 5.** Multivariate analysis of selected preoperative factors to predict the postoperative change of corrected distance visual acuity (ΔCDVA) after ICRS implantation

| Variables | Unstandardized Coefficients |  | Standardized Coefficients | p | Collinearity Statistics |  |
| --- | --- | --- | --- | --- | --- | --- |
|  | B | 95% CI | Beta |  | Tolerance | VIF |
| (Constant) | 0.589 | 0.269, 0.91 |  | <.001 |  |  |
| CDVAp | -0.713 | -0.809, -0.617 | -0.837 | <.001 | 0.727 | 1.375 |
| Rmin | -0.082 | -0.134, -0.030 | -0.169 | 0.002 | 0.808 | 1.238 |
| SPH | -0.010 | -0.017, -0.004 | -0.16 | 0.003 | 0.826 | 1.211 |

$R^2 = 0.541$ ; adjusted $R^2 = 0.534$ ; $F = 76.194$ ( $p < 0.001$ ); mean Cook's distance = $0.010 \pm 0.040$ ; Durbin-Watson test = 2.224
**Abbreviation:** CI, confidence interval; CDVAp, preoperative corrected distance visual acuity; SPH, spherical power; Rmin, minimum sagittal curvature; VIF, variance inflation factor

The predictability (R^2^) was 0.541, and the adjusted R^2^ was 0.534, with F = 76.194 (*p* < 0.001). No influential points or outliers were detected (mean Cook’s distance, 0.010 ± 0.040). The independence of the residuals (Durbin-Watson test = 2.224) and the lack of multicollinearity were confirmed. Patients with poorer preoperative visual acuity or higher CDVAp logMAR value demonstrated greater improvement in visual acuity after ring implantation. The model indicated that patients with more negative SPH (more myopic) and smaller Rmin would likely demonstrate poor visual gain when adjusted for CDVAp.

### 3.3 Validation of the predictive models

For the predictive model of ΔUDVA, the average absolute residual between predicted UDVA and actual UDVA was 0.294 ± 0.238, and the correct prediction was achieved only in 44% of cases within 0.22 logMAR.

For the predictive model of ΔCDVA, the average absolute residual between predicted CDVA and actual CDVA was 0.159 ± 0.159, and the correct prediction was achieved in 82% of cases within 0.22 logMAR

### 3.4 Complications of ICRS implantation

Both intraoperative and postoperative complications among the 351 eyes were recorded as secondary outcomes. There was one intraoperative complication in which a corneal perforation at the incision site using a stromal spreader was documented. This eye was successfully re-operated using a manual technique, approximately seven months following the first surgery. Postoperative complications included segment migration (17 eyes, 4.84%), ICRS-related infection (14 eyes, 3.99%), corneal melting (3 eyes, 0.85%), peri-annular deposits (2 eyes, 0.56%) and spontaneous fragmentation of the ring segment (1 eye, 0.28%). Twelve eyes underwent ring removal, four of which were because of infection-induced corneal melting. One patient requested ring explantation from his right eye owing to concerns about corneal irritation and a mild visual acuity drop post-surgery from CDVA 20/16 to 20/25.

## 4. DISCUSSION

According to our literature review, this may be the largest retrospective study of the relationship between ICRS-induced visual acuity changes and various demographic and preoperative factors of patients with keratoconus. The diversity of studied variable types was another strength of our study. This study was also the first attempt to create an equation to predict ΔUDVA following ICRS implantation. In the multiple linear regression analysis, UDVAp and PIavg seemed to be good predictors for ΔUDVA. However, this model had low predictability (Adjust R^2^ = 0.402) and low validity (accuracy rate less than 50%).

Regarding our univariate analysis, patients with poorer baseline visual acuity (larger value on the logMAR scale) were anticipated to demonstrate greater UDVA and CDVA improvement. This finding was consistent with those of previous studies [13, 15, 18, 21]. More myopic spherical power weakly correlated with more considerable CDVA gain (R = 0.123). This was in contrast to a study by Alio et al. [16], which found that patients with better visual acuity gain had less myopia than other groups. Several parameters derived from Scheimpflug-based tomography demonstrated significant correlations with ΔCDVA; however, only corneal astigmatism significantly correlated with ΔUDVA. Based on the Pearson coefficients, patients with more advanced baseline topography (steeper Km, K factor, and Kmax; and more prolate Q-value) tended to gain more lines of postoperative CDVA. However, our findings differed from previous studies, which found that patients with lower mean keratometry experienced more visual improvement [7, 16, 24].

The corneal thickness at the apex, thinnest point, and pupil center were significantly correlated with ΔCDVA in a similar direction, i.e., thinner corneal thickness tended to demonstrate more CDVA improvement. This finding contrasts that of Zare et al. [7], who stated that a patient with a thicker cornea (thinnest corneal thickness > 400 µm) would demonstrate greater improvements in UDVA and CDVA six months post-surgery. The relationships between ΔCDVA and all pachymetric indices were also shown in our study. ICRS-induced visual gain increased in patients with poorer pachymetric indices (higher preoperative BADD, higher PIavg, and lower ARTmax).

For topographic indices, higher ISV and CKI values and smaller Rmin significantly correlated with a more remarkable improvement in CDVA. The ISV is calculated from the standard deviation of individual sagittal radii from mean curvature. Higher ISV is an indicator of increased anterior corneal surface irregularity. CKI is the ratio between mean radius of curvature in the peripheral zone and mean radius of curvature in central zone, while Rmin denotes the maximum steepness of the cone [28]. As the CKI is higher, the cone of keratoconic cornea become far steeper, and Rmin usually become smaller. It can be inferred that the cornea with a more irregular surface and the steeper cone will gain a more favorable visual outcome from ICRS implantation. Nonetheless, a previous study had reported in the opposite direction that lower preoperative ISV and larger Rmin had a strong correlation with a better gain of UDVA and CDVA [24].

All aberrometry variables had a weak correlation with CDVA change in our study. The highly aberrated cornea was related to more gain of CDVA. However, many previous studies failed to demonstrate a significant correlation between aberrometry and visual acuity change after ICRS implantation [15, 22, 23].

Distances between the corneal apex, thinnest point, pupil center, and Kmax point were also studied for the hypothesis that longer distances might reflect more cone eccentricity and have more effect on visual acuity. However, this study failed to show any significant correlation between these distances and visual acuity change. Gatzioufas et al. also studied the association between visual outcome after ring implantation with distance from apex to thinnest point and from apex to maximum keratometry. They found that there was no statistically significant association [17].

In this study, the age at onset, age at surgery, history of rubbing or atopy, or intraocular pressure did not significantly correlate with ICRS-induced visual acuity change. Despite females experiencing more gain of UDVA than males in the univariate analysis, this association was disproven in the multivariate analysis, similar to previous results [19].

According to the result of univariate analysis in our study, CDVA improvement was anticipated to be better in more advanced keratoconus because it is usually associated with poorer baseline visual acuity, higher myopia, steeper keratometry, more prolateness, thinner pachymetry, higher surface irregularity, steeper cone, and higher aberration.

Our proposed model for predicting ΔCDVA included different predictors from those previously reported [21, 22]. Three variables, including CDVAp, SPH, and Rmin, were selected by the multivariate analysis in the predictive equation for ΔCDVA. This model corresponded to the findings of Pena-Garcia et al. [21] where CDVAp emerged as the most critical predictor of ΔCDVA at six months. While no keratometric parameters appeared in our model, the previously proposed model included at least one keratometric parameter [21, 22]. This may be attributed to different sets of potential variables in the multiple regression analysis and differences in ICRS manufacturers in each study. It could be noted that the sign of coefficients of SPH and Rmin had reversed from plus (in univariate analysis) to minus. This revealed that if two patients have equal CDVAp, one with less myopia and larger Rmin will be predicted to gain more CDVA improvement.

This study was limited by its sole focus on visual acuity at about six months post-surgery, as this time point appeared to be associated with short-term outcomes. Thus, further research with a longer follow-up is necessary to identify factors that predict the long-term effects of ICRS. Another limitation was a high rate of missing data due to the retrospective nature of this study. We excluded some cases because their medical records lacked pre-defined parameters; therefore, our results may not reflect real-world outcomes for all patients undergoing ICRS. The utility of the proposed models might be limited because its variables contained the spherical power from autorefraction, which is frequently unmeasurable in advanced keratoconus. Researchers are also interested in conducting studies on corneal biomechanical properties evaluated by the dynamic Scheimpflug analyzer (Corvis ST, Oculus, Wetzlar, Germany). This is based on the hypothesis that biomechanical properties of the cornea can be potential predictors for ΔCDVA following ICRS implantation [22]. Unfortunately, we could not perform the aforementioned study because a few patients underwent the dynamic Scheimpflug analysis before the ICRS surgery at our center. In conclusion, the baseline visual acuity, refractive error, and numerous parameters from Scheimpflug tomography can predict ICRS-induced changes in visual acuity six months post-surgery. These predictors, alongside the proposed model, could assist with appropriate case selection and correct timing for ICRS implantation. Considering these parameters may maximize visual gains for patients with keratoconus.

## Data Availability

All relevant data are within the manuscript and its Supporting Information files.

## ACKNOWLEDGEMENTS

We would like to thank Dollapas Punpanich, M.S., a consultant for statistical analysis, and Editage (www.editage.com) for English language editing.

## SUPPORTING INFORMATION

S1 File. The data spreadsheet (XLSX).

## REFERENCES

1. Zadnik K, Barr JT, Edrington TB, Everett DF, Jameson M, McMahon TT, et al. Baseline findings in the Collaborative Longitudinal Evaluation of Keratoconus (CLEK) Study. Invest Ophthalmol Vis Sci. 1998; 39(13):2537–46. Epub 1998/12/18. PubMed PMID: 9856763.

2. Wagner H, Barr JT, Zadnik K. Collaborative Longitudinal Evaluation of Keratoconus (CLEK) Study: methods and findings to date. Cont Lens Anterior Eye. 2007; 30(4):223–32. Epub 2007/05/08. doi: 10.1016/j.clae.2007.03.001. PubMed PMID: 17481941; PubMed Central PMCID: PMCPMC3966142.

3. Mohammadpour M, Heidari Z, Hashemi H. Updates on Managements for Keratoconus. J Curr Ophthalmol. 2018; 30(2):110–24. Epub 2018/07/11. doi: 10.1016/j.joco.2017.11.002. PubMed PMID: 29988906; PubMed Central PMCID: PMCPMC6034171.

4. Alfonso JF, Lisa C, Fernandez-Vega L, Madrid-Costa D, Montes-Mico R. Intrastromal corneal ring segment implantation in 219 keratoconic eyes at different stages. Graefes Arch Clin Exp Ophthalmol. 2011; 249(11):1705–12. Epub 2011/08/16. doi: 10.1007/s00417-011-1759-9. PubMed PMID: 21842130.

5. Shabayek MH, Alio JL. Intrastromal corneal ring segment implantation by femtosecond laser for keratoconus correction. Ophthalmology. 2007; 114(9):1643–52. Epub 2007/04/03. doi: 10.1016/j.ophtha.2006.11.033. PubMed PMID: 17400293.

6. Ferrara G, Torquetti L, Ferrara P, Merayo-Lloves J. Intrastromal corneal ring segments: visual outcomes from a large case series. Clin Exp Ophthalmol. 2012; 40(5):433–9. Epub 2011/09/10. doi: 10.1111/j.1442-9071.2011.02698.x. PubMed PMID: 21902789.

7. Zare MA, Hashemi H, Salari MR. Intracorneal ring segment implantation for the management of keratoconus: safety and efficacy. J Cataract Refract Surg. 2007; 33(11):1886–91. Epub 2007/10/30. doi: 10.1016/j.jcrs.2007.06.055. PubMed PMID: 17964393.

8. Chan SM, Khan HN. Reversibility and exchangeability of intrastromal corneal ring segments. J Cataract Refract Surg. 2002; 28(4):676–81. Epub 2002/04/17. doi: 10.1016/s0886-3350(01)01172-5. PubMed PMID: 11955910.

9. Alio JL, Shabayek MH, Artola A. Intracorneal ring segments for keratoconus correction: long-term follow-up. J Cataract Refract Surg. 2006; 32(6):978–85. Epub 2006/07/04. doi: 10.1016/j.jcrs.2006.02.044. PubMed PMID: 16814056.

10. Colin J, Malet FJ. Intacs for the correction of keratoconus: two-year follow-up. J Cataract Refract Surg. 2007; 33(1):69–74. Epub 2006/12/27. doi: 10.1016/j.jcrs.2006.08.057. PubMed PMID: 17189796.

11. Shetty R, Kurian M, Anand D, Mhaske P, Narayana KM, Shetty BK. Intacs in advanced keratoconus. Cornea. 2008; 27(9):1022–9. Epub 2008/09/25. doi: 10.1097/ICO.0b013e318172fc54. PubMed PMID: 18812766.

12. Carrasquillo KG, Rand J, Talamo JH. Intacs for keratoconus and post-LASIK ectasia: mechanical versus femtosecond laser-assisted channel creation. Cornea. 2007; 26(8):956–62. Epub 2007/08/28. doi: 10.1097/ICO.0b013e31811dfa66. PubMed PMID: 17721296.

13. Vega-Estrada A, Alio JL, Brenner LF, Javaloy J, Plaza Puche AB, Barraquer RI, et al. Outcome analysis of intracorneal ring segments for the treatment of keratoconus based on visual, refractive, and aberrometric impairment. Am J Ophthalmol. 2013; 155(3):575–84.e1. Epub 2012/12/12. doi: 10.1016/j.ajo.2012.08.020. PubMed PMID: 23218702.

14. Park J, Gritz DC. Evolution in the use of intrastromal corneal ring segments for corneal ectasia. Curr Opin Ophthalmol. 2013; 24(4):296–301. Epub 2013/05/15. doi: 10.1097/ICU.0b013e3283622a2c. PubMed PMID: 23665526.

15. Sedaghat MR, Momeni-Moghaddam H, Pinero DP, Akbarzadeh R, Moshirfar M, Bamdad S, et al. Predictors of Successful Outcome following Intrastromal Corneal Ring Segments Implantation. Curr Eye Res. 2019; 44(7):707–15. Epub 2019/03/15. doi: 10.1080/02713683.2019.1594945. PubMed PMID: 30868919.

16. Alio JL, Shabayek MH, Belda JI, Correas P, Feijoo ED. Analysis of results related to good and bad outcomes of Intacs implantation for keratoconus correction. J Cataract Refract Surg. 2006; 32(5):756–61. Epub 2006/06/13. doi: 10.1016/j.jcrs.2006.02.012. PubMed PMID: 16765791.

17. Gatzioufas Z, Panos GD, Elalfy M, Khine A, Hamada S, Lake D, et al. Effect of Conus Eccentricity on Visual Outcomes After Intracorneal Ring Segments Implantation in Keratoconus. J Refract Surg. 2018; 34(3):196–200. Epub 2018/03/10. doi: 10.3928/1081597x-20180115-02. PubMed PMID: 29522230.

18. Guyot C, Libeau L, Vabres B, Weber M, Lebranchu P, Orignac I. [Refractive outcome and prognostic factors for success of intracorneal ring segment implantation in keratoconus: A retrospective study of 75 eyes]. J Fr Ophtalmol. 2019; 42(2):118–26. Epub 2019/01/27. doi: 10.1016/j.jfo.2018.09.006. PubMed PMID: 30679126.

19. Jadidi K, Naderi M, Mosavi SA, Nejat F, Aghamolaei H, Serahati S. Pre-operative factors influencing post-operative outcomes from MyoRing implantation in keratoconus. Clin Exp Optom. 2019; 102(4):394–8. Epub 2018/12/12. doi: 10.1111/cxo.12859. PubMed PMID: 30536629.

20. Pena-Garcia P, Alio JL, Vega-Estrada A, Barraquer RI. Internal, corneal, and refractive astigmatism as prognostic factors for intrastromal corneal ring segment implantation in mild to moderate keratoconus. J Cataract Refract Surg. 2014; 40(10):1633–44. Epub 2014/09/30. doi: 10.1016/j.jcrs.2014.01.047. PubMed PMID: 25263039.

21. Pena-Garcia P, Vega-Estrada A, Barraquer RI, Burguera-Gimenez N, Alio JL. Intracorneal ring segment in keratoconus: a model to predict visual changes induced by the surgery. Invest Ophthalmol Vis Sci. 2012; 53(13):8447–57. Epub 2012/11/22. doi: 10.1167/iovs.12-10639. PubMed PMID: 23169881.

22. Piñero DP, Alio JL, Barraquer RI, Michael R. Corneal biomechanical changes after intracorneal ring segment implantation in keratoconus. Cornea. 2012; 31(5):491–9. Epub 2012/02/09. doi: 10.1097/ICO.0b013e31821ee9f4. PubMed PMID: 22314819.

23. Pinero DP, Alio JL, El Kady B, Coskunseven E, Morbelli H, Uceda-Montanes A, et al. Refractive and aberrometric outcomes of intracorneal ring segments for keratoconus: mechanical versus femtosecond-assisted procedures. Ophthalmology. 2009; 116(9):1675–87. Epub 2009/08/01. doi: 10.1016/j.ophtha.2009.05.016. PubMed PMID: 19643498.

24. Utine CA, Durmaz Engin C, Ayhan Z. Effects of Preoperative Topometric Indices on Visual Gain After Intracorneal Ring Segment Implantation For Keratoconus. Eye Contact Lens. 2018; 44 Suppl 2:S387–s91. Epub 2018/06/27. doi: 10.1097/icl.0000000000000490. PubMed PMID: 29944503.

25. Soto-Masías E, Galvez-Olortegui T, Galvez-Olortegui J, Iyo-Alberti F, Delgado-Becerra G. Factors associated with short-term visual improvement after intracorneal ring segments implantation. Revista Mexicana de Oftalmología (English Edition). 2020; 94. doi: 10.24875/RMOE.M20000116.

26. Sedaghat MR, Momeni-Moghaddam H, Belin MW, Piñero DP, Akbarzadeh R, Ambrósio R, Jr., et al. Comparative analysis of two different types of intracorneal implants in keratoconus: A corneal tomographic study. Eur J Ophthalmol. 2020:1120672120963449. Epub 2020/10/31. doi: 10.1177/1120672120963449. PubMed PMID: 33124461.

27. Neira W, Krootila K, Holopainen JM. Atopic dermatitis is a risk factor for intracorneal ring segment extrusion. Acta Ophthalmol. 2014; 92(6):e491–2. Epub 2014/04/15. doi: 10.1111/aos.12377. PubMed PMID: 24725421.

28. Kanellopoulos AJ, Asimellis G. Revisiting keratoconus diagnosis and progression classification based on evaluation of corneal asymmetry indices, derived from Scheimpflug imaging in keratoconic and suspect cases. Clin Ophthalmol. 2013; 7:1539–48. Epub 20130726. doi: 10.2147/opth.S44741. PubMed PMID: 23935360; PubMed Central PMCID: PMCPMC3735334.

